# Same-Session Clearance of Small Common Bile Duct Stones After Sphincterotomy Alone or With Adjunctive Extraction: A Single-Center Observational Study

**DOI:** 10.64898/2026.09.17.26363343

**Authors:** Fatemeh Yadollahzadeh Chari, Javad Shokri Shirvani, Seyed Hasan Abedi Valukalaei, Atefeh Yadollahzadeh Chari

## Abstract

**Background:** Small common bile duct stones may pass after endoscopic sphincterotomy, but leaving the duct without attempting extraction also leaves clearance uncertain. We compared same-session stone clearance after sphincterotomy alone with clearance after sphincterotomy followed by balloon-based and/or basket extraction.

**Methods:** We conducted a single-center observational study of 203 patients with common bile duct stones 5 mm or smaller who underwent therapeutic endoscopic retrograde cholangiopancreatography. The endoscopist chose the treatment during the procedure. Forty-seven patients underwent sphincterotomy alone, and 156 underwent sphincterotomy followed by balloon-based and/or basket extraction. The primary outcome was fluoroscopically confirmed clearance during the same procedure. Intraprocedural bleeding was a secondary outcome. We calculated unadjusted risk ratios and used Fisher’s exact test for sparse events.

**Results:** The mean age was 58.62 years (standard deviation 20.10), and 90 patients (44.3%) were men. Same-session clearance was documented in 21 of 47 patients (44.7%) after sphincterotomy alone and 130 of 156 (83.3%) after adjunctive extraction (risk ratio 1.87, 95% confidence interval 1.35 to 2.58; P < 0.001). A prominent ampulla was more common in the sphincterotomy-alone group (25.5% vs 10.9%; P = 0.012). Intraprocedural bleeding occurred in 2 patients after sphincterotomy alone and none after adjunctive extraction (4.3% vs 0%; Fisher’s exact P = 0.053).

**Conclusions:** Adjunctive extraction was associated with a higher rate of documented same-session clearance than sphincterotomy alone. Treatment was not randomized, the groups differed at baseline, and only two bleeding events occurred. These findings therefore do not prove that adjunctive extraction is causally superior or safer. Prospective studies using standardized techniques and follow-up are needed.

## Introduction

Endoscopic retrograde cholangiopancreatography (ERCP) is the standard endoscopic treatment for common bile duct stones. Endoscopic sphincterotomy (EST) opens the biliary outlet, and balloon or basket catheters can then be used to remove the stones. Guidelines recommend clearing the duct in patients who can tolerate treatment, while the choice of technique depends on stone size, duct anatomy, and local expertise [1–3].

For stones only a few millimeters in diameter, the best approach is less certain. Some may pass after sphincterotomy, making further instrumentation unnecessary. On the other hand, waiting for spontaneous passage leaves the duct uncleared at the end of the procedure and may lead to additional treatment. Because ERCP can cause pancreatitis, bleeding, infection, and perforation, any added maneuver should offer a clear clinical benefit [4,5].

Studies of papillary balloon dilation, limited sphincterotomy with dilation, and balloon or basket retrieval have reported high clearance rates, but the patients, stone sizes, and procedures have varied widely [6–11]. More recent cohort and randomized studies of stones up to 10 or 12 mm suggest that limited sphincterotomy with balloon dilation may reduce procedural burden without increasing early adverse events [12–14]. Evidence for stones 5 mm or smaller remains sparse.

We compared patients with common bile duct stones 5 mm or smaller who were treated with EST alone or with EST followed by adjunctive extraction. Our primary outcome was fluoroscopically confirmed clearance during the same ERCP session. Intraprocedural bleeding and use of mechanical lithotripsy were secondary outcomes.

## Methods

### Study Design and Setting

We conducted a single-center observational study in the endoscopy unit of Ayatollah Rouhani Hospital in Babol, Iran. Patients with clinical, laboratory, and imaging findings consistent with common bile duct stones were identified when they attended for therapeutic ERCP. Treatment was not randomized. The attending endoscopist chose the procedural strategy after considering the findings during ERCP and the patient’s clinical features. We prepared this report in accordance with the STROBE guidance for observational studies [16].

### Participants

Patients were eligible if imaging showed one or more common bile duct stones no larger than 5 mm. They also had to provide informed consent, have normal coagulation results before ERCP, not be taking anticoagulants, and have no history of ERCP or gastrointestinal surgery. We excluded patients when ERCP revealed a difficult stone, duct cannulation was difficult, uncontrolled bleeding or intestinal perforation occurred, or the patient withdrew consent.

### Procedures and Exposure Groups

All ERCP procedures were performed under fluoroscopic guidance, and every patient underwent EST. We classified patients according to the treatment recorded during the procedure: EST alone or EST followed by conventional extraction. Adjunctive extraction included balloon-based intervention, basket extraction, or both. We recorded mechanical lithotripsy and biliary stent placement separately. The treating endoscopist confirmed complete duct clearance fluoroscopically.

### Outcomes

The primary outcome was complete stone clearance during the index ERCP. The other recorded outcomes were planned observation for spontaneous passage and failure to clear the duct despite attempted extraction. Secondary procedural outcomes included mechanical lithotripsy, stent placement, and bleeding during the procedure. The dataset did not allow us to assess delayed bleeding, post-ERCP pancreatitis, cholangitis, later perforation, recurrent stones, or other outcomes requiring follow-up. Adverse events could not be graded with a standardized endoscopy lexicon [15].

### Statistical Analysis

Analyses were performed in SPSS version 22. We summarized continuous variables as means and standard deviations and compared groups with independent-samples t tests. Categorical variables are reported as counts and percentages and were compared with chi-square tests. We calculated unadjusted risk ratios and 95% confidence intervals from the reported 2-by-2 counts. Because the expected cell counts for intraprocedural bleeding were below 5, we used Fisher’s exact test for that comparison. Tests were two-sided, with P < 0.05 considered statistically significant. Individual-level data needed for an adjusted analysis were not available.

### Ethics

The Biomedical Research Ethics Committee of Babol University of Medical Sciences approved the study on 1 May 2021 (IR.MUBABOL.REC.1400.042). All participants gave informed consent. The analysis contained no directly identifiable patient information. Trial registration was not applicable because this was an observational study and participants were not prospectively assigned to an intervention.

## Results

### Participant Characteristics

We included 203 patients. Their mean age was 58.62 years (standard deviation 20.10); 90 patients (44.3%) were men and 113 (55.7%) were women. Fifty-one patients (25.1%) had previously undergone cholecystectomy. Nineteen (9.4%) presented with acute cholangitis and 6 (3.0%) with acute pancreatitis. Table 1 summarizes the cohort.

**Table 1.** Participant characteristics.

| Characteristic | Overall cohort N = 203 |
| --- | --- |
| Age in years mean SD | 58.62 20.10 |
| Male sex | 90 44.3 |
| Female sex | 113 55.7 |
| Hypertension | 19 9.4 |
| Diabetes mellitus | 8 3.9 |
| Ischemic heart disease | 10 4.9 |
| Previous cholecystectomy | 51 25.1 |
| Opioid use | 24 11.8 |
| Smoking | 14 6.9 |
| Acute cholangitis at presentation | 19 9.4 |
| Acute pancreatitis at presentation | 6 3.0 |

Values are n percent unless otherwise stated. SD standard deviation.

### Procedural Findings

The mean stone diameter was 4.74 mm (standard deviation 0.81). Twenty-three patients (11.3%) had a periampullary diverticulum, 29 (14.3%) had a prominent ampulla, and 34 (16.7%) had a dilated bile duct. Cannulation method was recorded for 196 patients: 163 underwent standard cannulation and 33 underwent precut cannulation. Balloon-based intervention was used in 103 patients and a basket in 110; some patients received both. Twelve patients underwent mechanical lithotripsy, and 3 received a biliary stent.

The duct was cleared during the index procedure in 151 patients (74.4%). The endoscopist chose observation for expected spontaneous passage in 42 patients (20.7%), and attempted extraction failed in 10 (4.9%). Two bleeding events occurred during ERCP. Table 2 summarizes the procedural findings and outcomes.

**Table 2.**
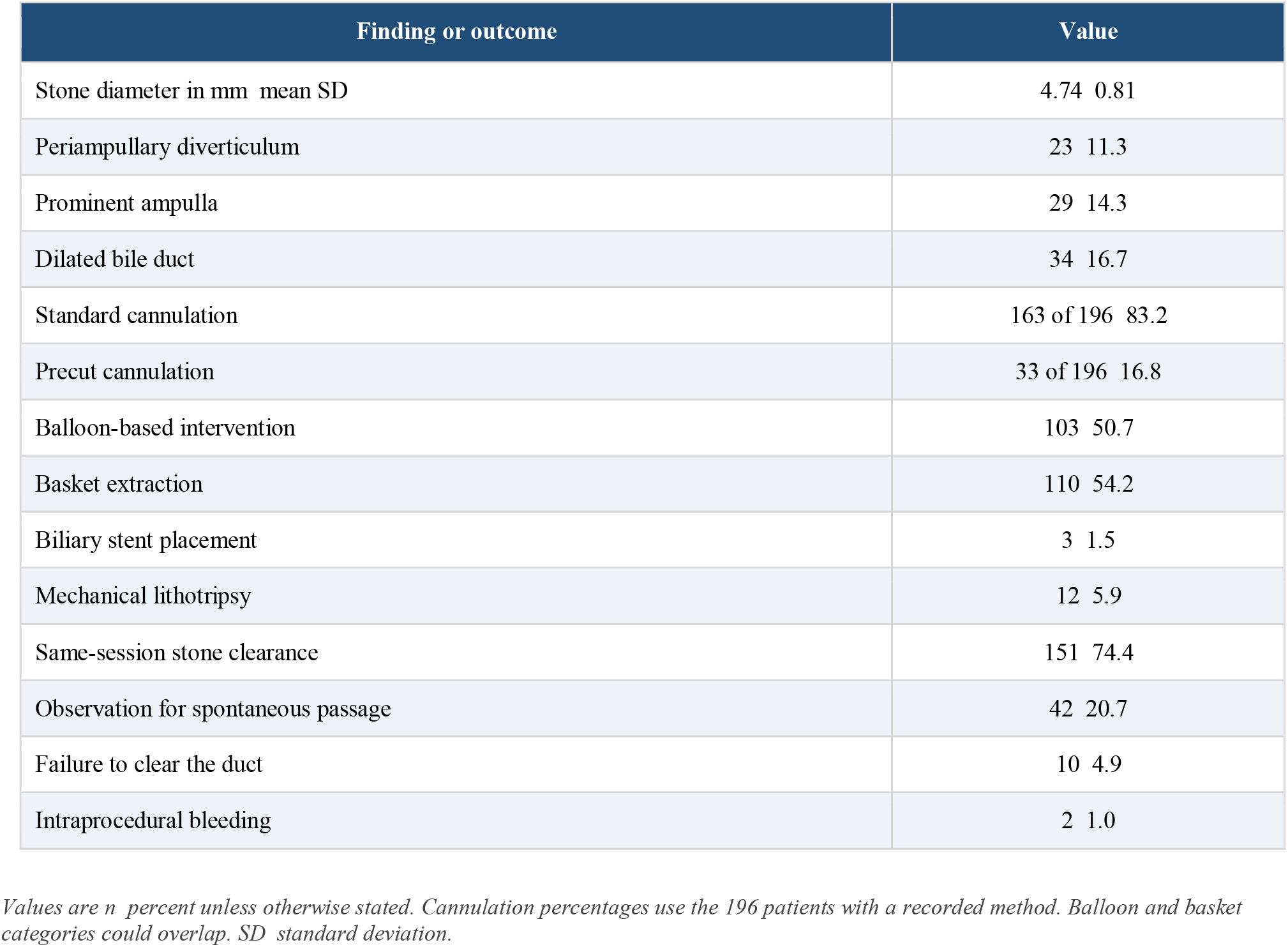
Procedural findings and outcomes.

### Comparison of Treatment Groups

Forty-seven patients (23.2%) underwent EST alone, while 156 (76.8%) underwent EST followed by adjunctive extraction. Patients in the adjunctive-extraction group were older on average (60.56 vs 52.17 years; P = 0.013). A prominent ampulla was less common in this group (17 of 156 [10.9%] vs 12 of 47 [25.5%]; P = 0.012). None of the other recorded baseline comparisons reached statistical significance (all P > 0.05).

Same-session clearance was documented in 130 of 156 patients (83.3%) after adjunctive extraction and 21 of 47 (44.7%) after EST alone. The unadjusted probability of clearance was 1.87 times as high with adjunctive extraction (95% confidence interval 1.35 to 2.58; P < 0.001). Bleeding during the procedure occurred in 2 of 47 patients (4.3%) after EST alone and in none of the 156 patients after adjunctive extraction. This difference did not reach conventional statistical significance with Fisher’s exact test (P = 0.053). The absolute risk difference was −4.3 percentage points (approximate 95% confidence interval −10.0 to 1.5).

**Table 3.** Main comparison of treatment groups.

| Outcome | EST alone n = 47 | EST plus adjunctive extraction n = 156 | Effect estimate | P value |
| --- | --- | --- | --- | --- |
| Age in years mean | 52.17 | 60.56 | +8.39 years | 0.013 |
| Prominent ampulla | 12 25.5 | 17 10.9 | RR 0.43 0.22 to 0.83 | 0.012 |
| Same-session clearance | 21 44.7 | 130 83.3 | RR 1.87 1.35 to 2.58 | < 0.001 |
| Intraprocedural bleeding | 2 4.3 | 0 0 | RD -4.3 points -10.0 to 1.5 | 0.053 |

Values are n percent unless otherwise stated. Effects compare adjunctive extraction with EST alone; ranges are 95 percent confidence intervals. Bleeding uses Fisher exact testing; other P values use t or chi-square tests. EST endoscopic sphincterotomy. RD risk difference. RR risk ratio.

## Discussion

### Principal Findings

In this cohort of patients with common bile duct stones 5 mm or smaller, EST followed by adjunctive extraction was associated with more frequent clearance during the index ERCP than EST alone. The absolute difference was 38.6 percentage points, and the unadjusted risk ratio was 1.87. In practical terms, active extraction made it more likely that the endoscopist could confirm an empty duct before the procedure ended.

Only two bleeding events occurred, both in the EST-alone group. Because the expected cell counts were small, Fisher’s exact test was more appropriate than a chi-square test; the resulting P value was 0.053. The point estimate favored adjunctive extraction, but the confidence interval was wide. The study therefore cannot establish a difference in bleeding risk.

### Comparison With Previous Evidence

Our clearance findings are consistent with earlier comparisons of sphincterotomy plus balloon dilation and sphincterotomy alone. Ishii and colleagues reported fewer additional ERCP sessions after minimal EST with papillary balloon dilation, while a meta-analysis by Dong and colleagues found higher first-session clearance and less use of mechanical lithotripsy [7,8]. More recent randomized and propensity-adjusted studies have also reported procedural advantages for limited EST combined with papillary balloon dilation in stones up to 10 or 12 mm [12–14].

Those studies are not directly comparable with ours. The adjunctive group in our cohort included a mixture of balloon-based and basket maneuvers rather than a single, standardized sphincterotomy-plus-balloon-dilation protocol. The retrieval device itself may also affect clearance and procedure time [11]. Our results should therefore be read as a comparison between an EST-alone strategy and a strategy that added active extraction, not as a definitive test of any one dilation technique.

### Clinical Interpretation

Confirming clearance during ERCP is useful because it completes treatment and reduces uncertainty about retained stones. Still, part of the difference we observed is built into the treatment strategies: one group received a device intended to remove the stone, whereas some patients in the EST-alone group were deliberately observed for spontaneous passage. The study shows that adjunctive extraction improves immediate confirmation of clearance; it does not show that every patient with a stone 5 mm or smaller needs additional instrumentation.

The endoscopist’s treatment choice may also have influenced the result. The groups differed in age and ampullary anatomy, and unmeasured clinical features may have affected which treatment a patient received. We could not perform multivariable or propensity-score adjustment without individual-level data. The association should not therefore be interpreted as proof that adjunctive extraction caused the higher clearance rate.

### Strengths and Limitations

This study addressed a focused question in a cohort restricted to stones 5 mm or smaller. Stone size and procedural outcomes were recorded during ERCP, and the endoscopist verified the primary outcome fluoroscopically. Using an exact test for the rare bleeding events provides a more reliable account of the uncertainty around that result.

The main limitations are the observational, single-center, nonrandomized design and the absence of an adjusted analysis. Balloon and basket use overlapped, so we cannot separate the effects of individual techniques. We also lacked procedure duration, device specifications, balloon diameter and inflation time, operator experience, and complete follow-up after ERCP. Cannulation method was missing for 7 patients. Bleeding was recorded only during the procedure, and the study did not adequately capture delayed bleeding, post-ERCP pancreatitis, cholangitis, later perforation, recurrent stones, or repeat procedures. These gaps prevent firm conclusions about comparative safety and long-term effectiveness.

## Conclusions

Among patients with common bile duct stones 5 mm or smaller, EST followed by adjunctive extraction was associated with more frequent fluoroscopically confirmed clearance during the index ERCP than EST alone. The two bleeding events were too few to support a reliable conclusion about safety. Future prospective studies should use a standardized extraction technique, prespecified adverse-event definitions, complete follow-up, and either randomization or adequate adjustment for treatment selection.

## Data Availability

All aggregate data supporting the findings of this study are contained in the manuscript. Individual-level participant data are not publicly available because of ethics, privacy, and institutional restrictions.

## Declarations

### Ethics Approval and Consent to Participate

The Biomedical Research Ethics Committee of Babol University of Medical Sciences approved the study on 1 May 2021 (IR.MUBABOL.REC.1400.042). All participants gave informed consent.

### Trial Registration

Not applicable. This was an observational study, and participants were not prospectively assigned to an intervention.

### Consent for Publication

Not applicable. The manuscript contains no identifiable individual patient information.

### Funding

No external funding was reported in the thesis or the previous manuscript draft.

### Competing Interests

No competing interests were reported in the thesis or the previous manuscript draft.

### Data Availability

All aggregate data used in this analysis are reported in the article. Requests for access to deidentified individual-level data may be directed to the corresponding author and would be subject to applicable ethics, privacy, and institutional requirements.

### Reporting Guideline

This report was prepared in accordance with the STROBE guidance for observational studies.

## Notes

### Competing Interest Statement

The authors have declared no competing interest.

### Author Declarations

The Biomedical Research Ethics Committee of Babol University of Medical Sciences gave ethical approval for this work (approval number IR.MUBABOL.REC.1400.042; 1 May 2021).

